# The distribution and spread of susceptible and resistant *Neisseria gonorrhoeae* across demographic groups in a major metropolitan center

**DOI:** 10.1101/2020.04.30.20086413

**Authors:** Tatum D. Mortimer, Preeti Pathela, Addie Crawley, Jennifer L. Rakeman, Ying Lin, Simon R. Harris, Susan Blank, Julia A. Schillinger, Yonatan H. Grad

## Abstract

**Background:** Genomic epidemiology studies of gonorrhea in the United States have primarily focused on national surveillance for antibiotic resistance, and patterns of local transmission between demographic groups of resistant and susceptible strains are unknown.

**Methods:** We analyzed a convenience sample of genome sequences, antibiotic susceptibility, and patient data from 897 gonococcal isolates cultured at the NYC Public Health Laboratory from NYC Department of Health and Mental Hygiene (DOHMH) Sexual Health Clinic (SHC) patients, primarily in 2012-13. We reconstructed the gonococcal phylogeny, defined transmission clusters using a 10 non-recombinant single nucleotide polymorphism threshold, tested for clustering of demographic groups, and placed NYC isolates in a global phylogenetic context.

**Results:** The NYC gonococcal phylogeny reflected global diversity with isolates from 22/23 of the prevalent global lineages (96%). Isolates clustered on the phylogeny by patient sexual behavior (p<0.001) and race/ethnicity (p<0.001).Minimum inhibitory concentrations were higher across antibiotics in isolates from men who have sex with men compared to heterosexuals (p<0.001) and white heterosexuals compared to black heterosexuals (p<0.01). In our dataset, all large transmission clusters (≥10 samples) of *N. gonorrhoeae* were susceptible to ciprofloxacin, ceftriaxone, and azithromycin and comprised isolates from patients across demographic groups.

**Conclusions:** All large transmission clusters were susceptible to gonorrhea therapies, suggesting that resistance to empiric therapy was not a main driver of spread, even as risk for resistance varied across demographic groups. Further study of local transmission networks is needed to identify drivers of transmission.

## Introduction

Rates of reported *Neisseria gonorrhoeae* infections (gonorrhea) in the United States have fluctuated in the antibiotic era. After peaking in the early 1970s, rates dropped to a historic low in the US in 2009 [1]. However, since then, rates have climbed, with 583,405 gonorrhea cases reported in 2018, up 82.6% from 2009 [1]. The trends in reported cases of gonorrhea vary by sexual behavior and demographics. Men who have sex with men (MSM) have higher rates of gonorrhea infection than heterosexual people, and rates in the MSM population have been increasing more rapidly [1]. Men and women aged 20-24 years old are at highest risk, and there are major disparities in gonorrhea infection rates across race/ethnicity subgroups; rates among black Americans were 7.7 fold higher than rates among white Americans 2018 [1].

*N. gonorrhoeae* infection has become a major public health concern [2,3], given its increasing incidence and decreased antibiotic susceptibility [4–6]. As gonorrhea treatment is most commonly empiric, treatment recommendations in the US are informed by antibiotic susceptibility patterns of isolates cultured at public health laboratories from specimens collected at sentinel sexually transmitted disease clinics [1].The prevalence of resistance varies by risk group and has resulted at times in population-specific empiric antibiotic treatment recommendations [7]. However, the extent to which gonorrhea spreads between demographic groups has been unclear. Understanding the patterns of spread and the factors driving change in incidence is critical for the design of effective clinical and public health measures to reduce the overall burden of disease and slow the spread of resistance.

Genomic epidemiology studies have begun to address questions on transmission between demographic groups as well as on resistance prevalence. Analysis of a global collection of *N. gonorrhoeae* spanning several decades revealed two major circulating gonococcal lineages, a lineage that is primarily associated with MSM and that tends to be multidrug resistant and another associated with heterosexuals that tends to be more antibiotic susceptible [8]. However, studies of transmission within smaller geographic regions and focused on more recent samples demonstrated extensive bridging between MSM and heterosexual populations, suggesting limited gonococcal lineage association with subpopulations [9,10]. Studies in the United States have focused primarily on national samples of strains resistant to antibiotics to define the genetic determinants of antibiotic resistance [11–13] or transmission of specific resistant lineages [14], and sampling may impact our understanding of gonorrhea transmission using genomic methods.

We sought to understand the local patterns of transmission of gonorrhea and antibiotic resistance across demographic groups. To do so, we sequenced and analyzed genomes from a sample of *N. gonorrhoeae* isolates cultured at the New York City (NYC) Public Health Laboratory (PHL) from diagnostic specimens collected from individuals attending Sexual Health Clinics (SHCs) run by the NYC Department of Health and Mental Hygiene (DOHMH). We used genomic data to describe transmission of antibiotic resistant and sensitive gonorrhea in NYC, and, using detailed patient demographic and clinical information linked to the isolates, we examined the relationship between patient groups, gonorrhea transmission, and antibiotic resistance.

## Methods

### Sample collection

The retrospective convenience sample of 897 isolates of *N. gonorrhoeae* were cultured by NYC PHL from specimens collected from 822 patients who visited the NYC DOHMH SHCs between July 2011 and September 2015. The majority of isolates (98.4%) were collected between January 2012 and June 2014. The isolates were collected as part of NYC’s standard contribution of urethral isolates to the Gonococcal Isolate Surveillance Project (GISP) and routine culture for clinical care, e.g., test of cure after treatment with a non-recommended regimen, persistent symptoms, or indications for anorectal or oropharyngeal testing prior to the availability of anorectal and oropharyngeal nucleic acid amplification test (NAAT) in SHCs.

### Demographic data collection

The following SHC visit-level data on patients whose isolates were included in the sample were extracted from SHC electronic medical records: gender, age, race/ethnicity, gender of sex partners, anatomic site of specimen collection, and HIV status (Supplementary Table 1).

### Representativeness of sampled isolates

Our sample was derived from cultured isolates stored by the NYC PHL. Using χ^2^ tests, we compared the characteristics (race/ethnicity, age, gender, sex of partners) of patients with study isolates to those of patients with gonorrhea detected by NAAT-only at NYC SHCs during the same time period. Patients contributing isolates to the study were excluded from the NAAT-positive comparison group. We considered patients with gonorrhea detected at multiple anatomic sites on the same day to have a single gonorrhea diagnosis; a patient could be counted more than once if they were diagnosed with gonorrhea more than 30 days after an earlier gonorrhea diagnosis.

### Isolate growth, DNA extraction, and whole genome sequencing

Previously stored *N. gonorrhoeae* isolates were subcultured at the NYC PHL using Chocolate II Agar plates (BBL) in a humidified 5% CO_2_ incubator at 35°C. DNA was extracted from fresh overnight subculture from single colonies using QIAcube with QIAamp DNA Mini Kit (Qiagen), and the quality of extracted DNA was checked with NanoDrop (Thermo Scientific). Whole genome sequencing (WGS) was performed on the Illumina HiSeq using standard protocols at the Wellcome Sanger Institute.

### Antibiotic susceptibility testing

Azithromycin (AZM), cefixime (CFM), ceftriaxone (CRO), and ciprofloxacin (CIP) E-strips (bioMérieux) were used to determine minimum inhibitory concentrations (MIC) at NYC PHL at the time that the initial clinical culture was performed. The CDC alert values for reduced susceptibility to these antibiotics are AZM ≥ 2 μg/mL, CFM ≥ 0.25 μg/mL, CRO ≥ 0.125 μg/mL; only ciprofloxacin has a defined resistance breakpoint per CLSI, which is CIP ≥ 1 μg/mL [1,15,16].

### Quality control of genomic data

FastQC [17] was used to assess the quality of WGS reads. Samples were removed if quality scores were poor across reads or if the total number of reads was not sufficient to cover the expected size of the *N. gonorrhoeae* genome (~2Mb). Metaphlan 2.5.0 [18] was used to identify sequences of non-gonococcal origin, which were removed from the dataset.

### Assembly

Spades 3.12 [19] was used for *de novo* assembly. Assemblies were corrected using the –careful option, and contigs with less than 10X coverage or 500 nucleotides in length were filtered. Assemblies were annotated with Prokka 1.13 [20]. Additionally, reads from all isolates were mapped to NCCP11945 (NC_011035.1) [21] using BWA-MEM [22]. Pilon v 1.16 was used to identify variants with a minimum depth of 10 reads and minimum mapping quality of 20 [23]. Pseudogenomes were created by incorporating single nucleotide polymorphisms (SNPs), small deletions, and uncertain positions into the reference genome.

### Phylogenetic analysis and clustering

We used Gubbins [24] to identify and mask recombinant regions and RAxML [25] to estimate the phylogeny of our sample. The phylogeny was based on 27112 non-recombinant SNPs; the total number of SNPs from the unmasked alignment was 63338. We used fastbaps [26] to partition the phylogeny into BAPS groups. To identify transmission clusters within our sample, we grouped isolates based on non-recombinant SNP differences (Supplementary Material, Supplementary Figure 1).

### Visualizations

Phylogenies were visualized with ITOL [27]. Plots were made with ggplot [28], ggpubr (https://rpkgs.datanovia.com/ggpubr/), and cowplot (https://github.com/wilkelab/cowplot).

### Statistics

Fritz and Purvis D [29], implemented in caper (https://cran.r-project.org/web/packages/caper/index.html), was used to test for phylogenetic structure of discrete traits. Fisher’s Exact Test or χ^2^ test was used to test for associations between categorical variables. The Kruskal Wallis test was used to test for associations between MICs and patient demographics, and the Wilcoxon rank sum test was used to test the significance of pairwise comparisons.

### Data availability

WGS data were deposited in the European Nucleotide Archive (ERA) under study accession PRJEB10016 (Supplementary Table 1). All additional data and scripts are available at https://github.com/gradlab/GC_NYC.

The NYC DOHMH considered this project to be public health surveillance that does not meet the Office of Human Research Protections definition of human subjects research.

## Results

In our convenience sample of isolates, most were cultured from specimens collected from male patients (95.2%, 854/897), and of those, 73.1% (613/854) were isolated from specimens collected from men who have sex with men (MSM; Table 1). Isolates cultured from specimens from heterosexuals were primarily collected from black patients (81.4%, 180/221), whereas those from MSM were more distributed among races/ethnicities (40.0%, 245/613 Non-Hispanic (NH)-White, 27.2%, 167/613 NH-Black, 23.5%, 144/613 Hispanic, 4.7%, 29/613 NH-Asian, and 4.6%, 28/613 NH-Other). *N. gonorrhoeae* was isolated from urethral (82.0%, 700/854), rectal (14.8%, 126/854), and pharyngeal (3.3%, 28/854) specimens in men. In women, *N. gonorrhoeae* was isolated from cervical (92.0%, 34/37), pharyngeal (5.4%, 2/37), and rectal (2.7%, 1/37) specimens.

**Table 1.**
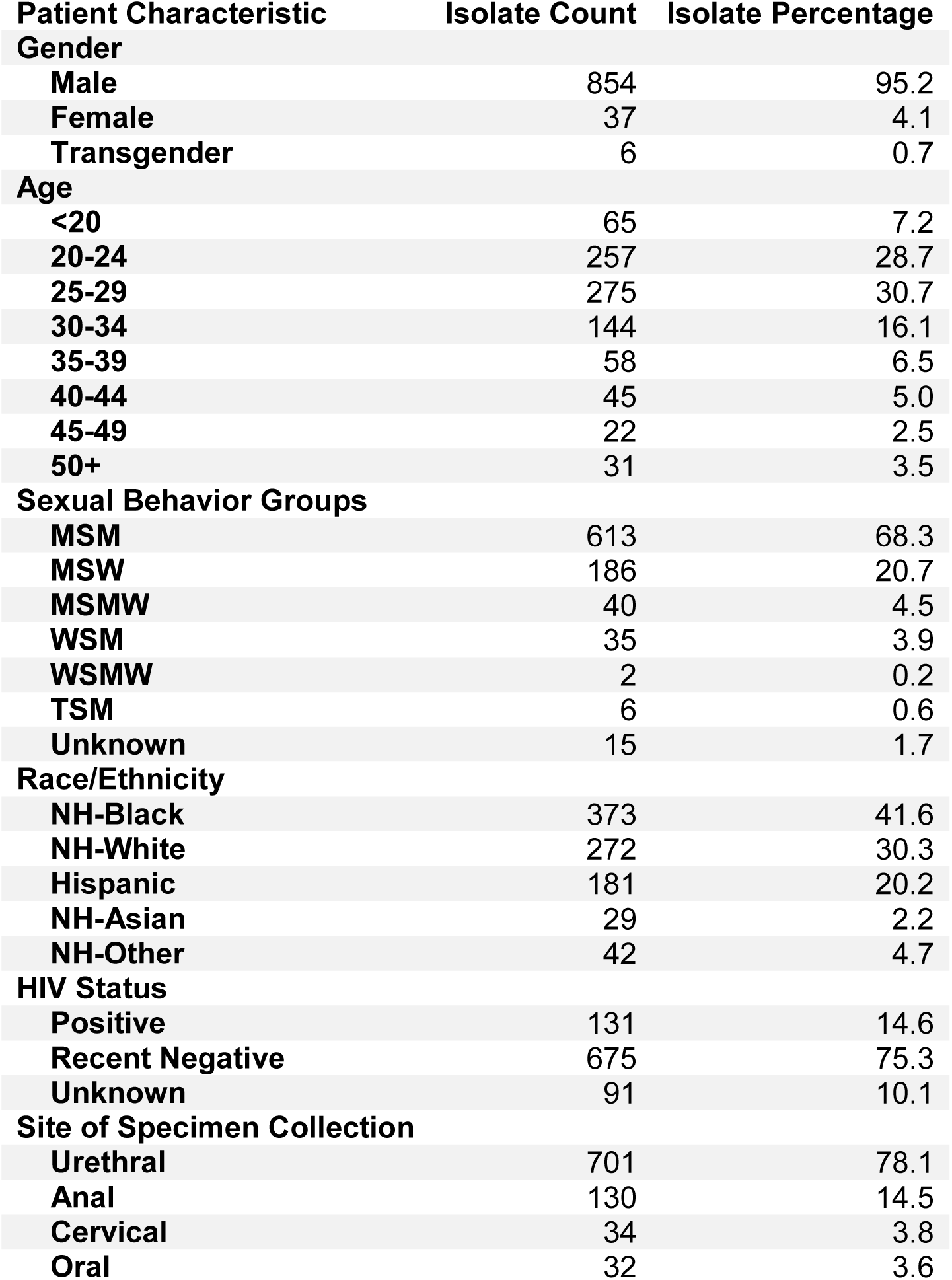
Patient demographics associated with *Neisseria gonorrhoeae* isolates from people attending New York City Sexual Health Clinics, 2011-2015.

| Patient Characteristic | Isolate Count | Isolate Percentage |
| --- | --- | --- |
| **Gender** |  |  |
| Male | 854 | 95.2 |
| Female | 37 | 4.1 |
| Transgender | 6 | 0.7 |
| **Age** |  |  |
| <20 | 65 | 7.2 |
| 20-24 | 257 | 28.7 |
| 25-29 | 275 | 30.7 |
| 30-34 | 144 | 16.1 |
| 35-39 | 58 | 6.5 |
| 40-44 | 45 | 5.0 |
| 45-49 | 22 | 2.5 |
| 50+ | 31 | 3.5 |
| **Sexual Behavior Groups** |  |  |
| MSM | 613 | 68.3 |
| MSW | 186 | 20.7 |
| MSMW | 40 | 4.5 |
| WSM | 35 | 3.9 |
| WSMW | 2 | 0.2 |
| TSM | 6 | 0.6 |
| Unknown | 15 | 1.7 |
| **Race/Ethnicity** |  |  |
| NH-Black | 373 | 41.6 |
| NH-White | 272 | 30.3 |
| Hispanic | 181 | 20.2 |
| NH-Asian | 29 | 2.2 |
| NH-Other | 42 | 4.7 |
| **HIV Status** |  |  |
| Positive | 131 | 14.6 |
| Recent Negative | 675 | 75.3 |
| Unknown | 91 | 10.1 |
| **Site of Specimen Collection** |  |  |
| Urethral | 701 | 78.1 |
| Anal | 130 | 14.5 |
| Cervical | 34 | 3.8 |
| Oral | 32 | 3.6 |

During the study period, 59,420 gonorrhea infections in 48,648 people living in NYC were reported to the DOHMH. Eighteen percent of gonorrhea infections in NYC and 23% of infections among men in NYC are in patients seen at the DOHMH SHCs. The vast majority of *N. gonorrhoeae* infections diagnosed in the SHCs were detected by NAAT during the period our isolates were collected (9740 infections excluding patients contributing to study isolates). People less than 20 years old, heterosexuals, and non-white MSM were underrepresented in the population contributing study isolates as compared to the population with NAAT-positive events at the DOHMH SHC.

For 33 people, *N. gonorrhoeae* was isolated from specimens collected from more than one anatomic site at the same visit. In 18.2% (6/33) of these patients, the isolates from different anatomic sites were distinct strains, which we defined as isolates with greater than ten non-recombinant SNP differences (Supplementary Figure 2A). We also observed 87 isolates from 41 people who returned to the SHCs multiple times; 90.5% (38/42) of re-infections were with a new strain. Re-infection with the same strain occurred up to 223 days after the original collection date (Supplementary Figure 2B). For two pairs of isolates, one from specimens collected at the same visit and one from specimens collected at multiple visits, the SNP distances were just outside our threshold for considering the isolates the same strain (12 and 15 SNPs). One isolate from each pair has unique recombination events, so SNP distances in these pairs may be inflated by unidentified recombination.

### N. gonorrhoeae *population structure*

We reconstructed the population structure of our NYC sample to examine the relationship between the gonococcus population and patient demographic groups. In a maximum likelihood phylogeny based on the non-recombinant portions of the genome, we observed two main lineages of *N. gonorrhoeae* (Figure 1A). Lineage A was significantly associated with MSM isolates, and lineage B was significantly associated with isolates from heterosexual patients (p < 2.2 × 10^−16^, Figure 1B). The phylogeny was significantly structured by sexual behavior (p < 0.001) and race/ethnicity (p < 0.001).

To test whether the gonococcal population in NYC represented a subset of globally circulating *N. gonorrhoeae*, we compiled a dataset of publicly available *N. gonorrhoeae* genomes [30] (Supplementary Table 2) and described the population structure using fastbaps [26]. The genome of at least one NYC isolate was present in 72% (33/46) of BAPS groups. We defined common BAPS groups as those containing at least one percent of the total genomes in the global dataset. A NYC genome was present in 96% (22/23) of these common groups.

**Figure 1.**
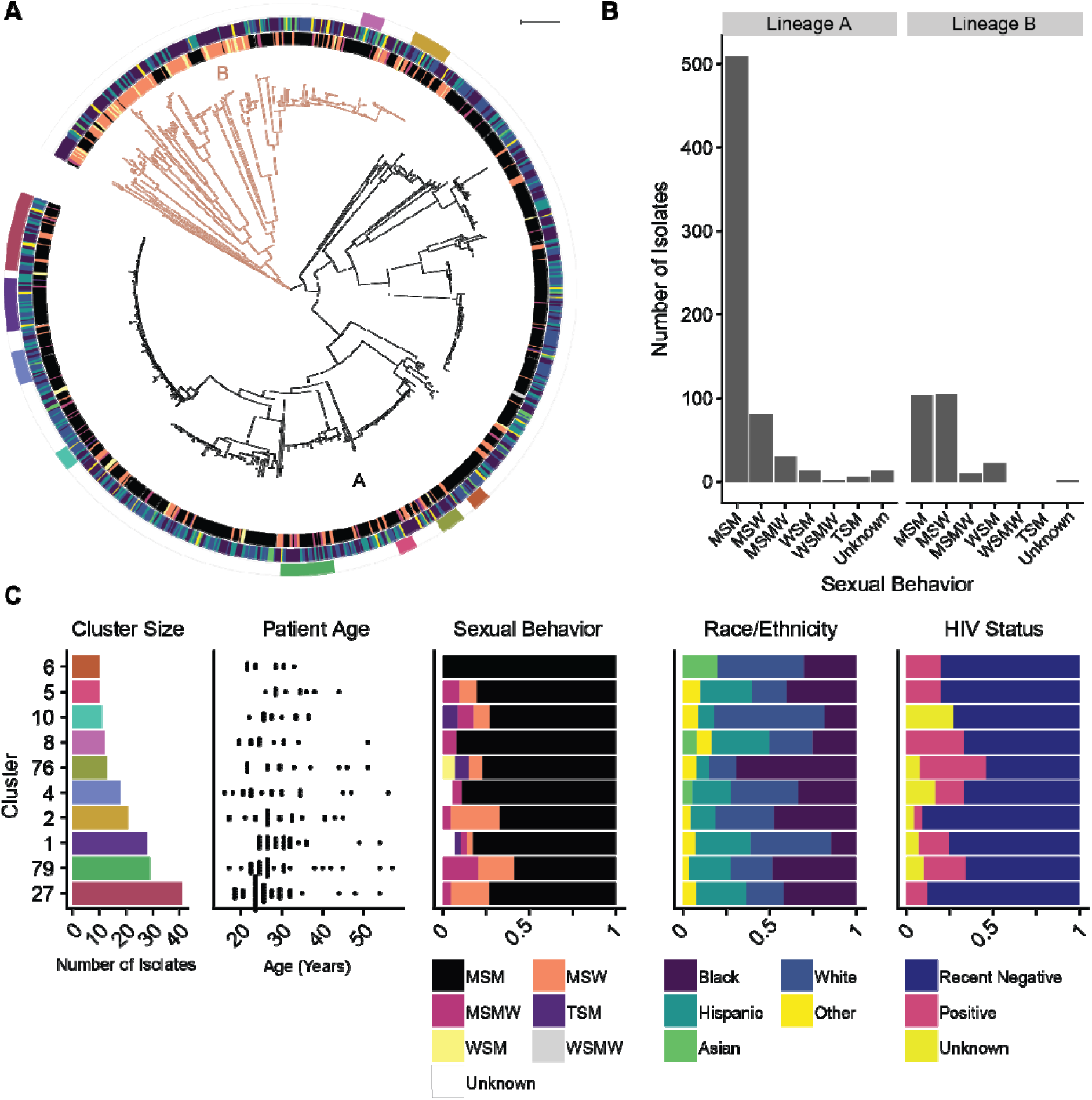
Population structure and transmission clusters of *Neisseria gonorrhoeae* in New York City. **A)** *N. gonorrhoeae* phylogeny is structured by sexual behavior groups, defined by patient gender and the sex of their sex partners, and race/ethnicity. The maximum likelihood phylogeny was estimated from a pseudogenome alignment with recombinant regions masked. The phylogeny can be divided into two main lineages, lineage A (black) and lineage B (orange). The annotation rings represent sexual behavior groups, race/ethnicity, and transmission clusters with >= 10 isolates from the innermost to outermost rings. For the sexual behavior groups, black is men who have sex with men (MSM), orange is men who have sex with women (MSW), pink is men who sex with men and women (MSMW), yellow is women who have sex with women (WSM), gray is women who have sex with men and women (WSMW), purple is transgender persons who have sex with men (TSM), and white is unknown sexual behavior group. For the race/ethnicity annotation, purple is black, blue is white, teal is Hispanic, green is Asian, and yellow is other. B) Lineage A is associated with MSM, and lineage B is associated with heterosexual patients. We found a significant association between the major lineages and sexual behavior group (p p < 2.2 × 10-16). **C)** Strains comprising largest transmission clusters are transmitting across demographic groups. Transmission connections were identified using a 10 non-recombinant SNP cutoff, and transmission clusters were defined as clusters of isolates connected to at least one other member of the cluster and any additional isolates nested within the phylogeny. Using this method, we identified 10 clusters with at least 10 isolates (leftmost graph, colors match annotation ring in panel A). Strains are transmitting across age groups; each point in the age graph (second to left) is the age of a patient associated with an isolate in the cluster. Strains are also transmitting across MSM and heterosexual networks and multiple races/ethnicities (third and fourth to left); color blocks correspond to the proportion of isolates within the cluster associated with each group (legend below graph).

### Characteristics of transmission clusters

We found that 65% (581/897) of isolates were clustered with at least one other isolate in our sample and identified 10 clusters with at least 10 isolates per cluster (Figure 1A,C). Using patient metadata, we characterized the demographic variation among the largest clusters (≥10 isolates) in our dataset (Figure 1C). Seven of ten clusters contained isolates from both MSM and heterosexuals, suggesting bridging across sexual networks. The individuals with gonococcal isolates in these large clusters also represented multiple races/ethnicities and varied age. The distribution of HIV status across the phylogeny was not significantly different from random (p = 0.28), and the transmission clusters included HIV positive and negative patients (Figure 1C).

### Characteristics of unclustered isolates

We compared the demographic characteristics of clustered and unclustered isolates to determine whether unsampled transmission was associated with particular patient groups. MSM associated isolates were more likely to be clustered than isolates from heterosexual patients (p = 5.0 × 10^−4^, Figure 2AB). We examined the association between race/ethnicity and clustering of isolates separately for each sexual behavior group. We found no association between race/ethnicity and clustering among genomes from isolates from MSM (p = 0.518). However, among heterosexuals, race/ethnicity was associated with clustering (p = 0.046, Figure 3A); genomes from isolates from white patients were more likely to be unclustered (Figure 3B).

**Figure 2.**
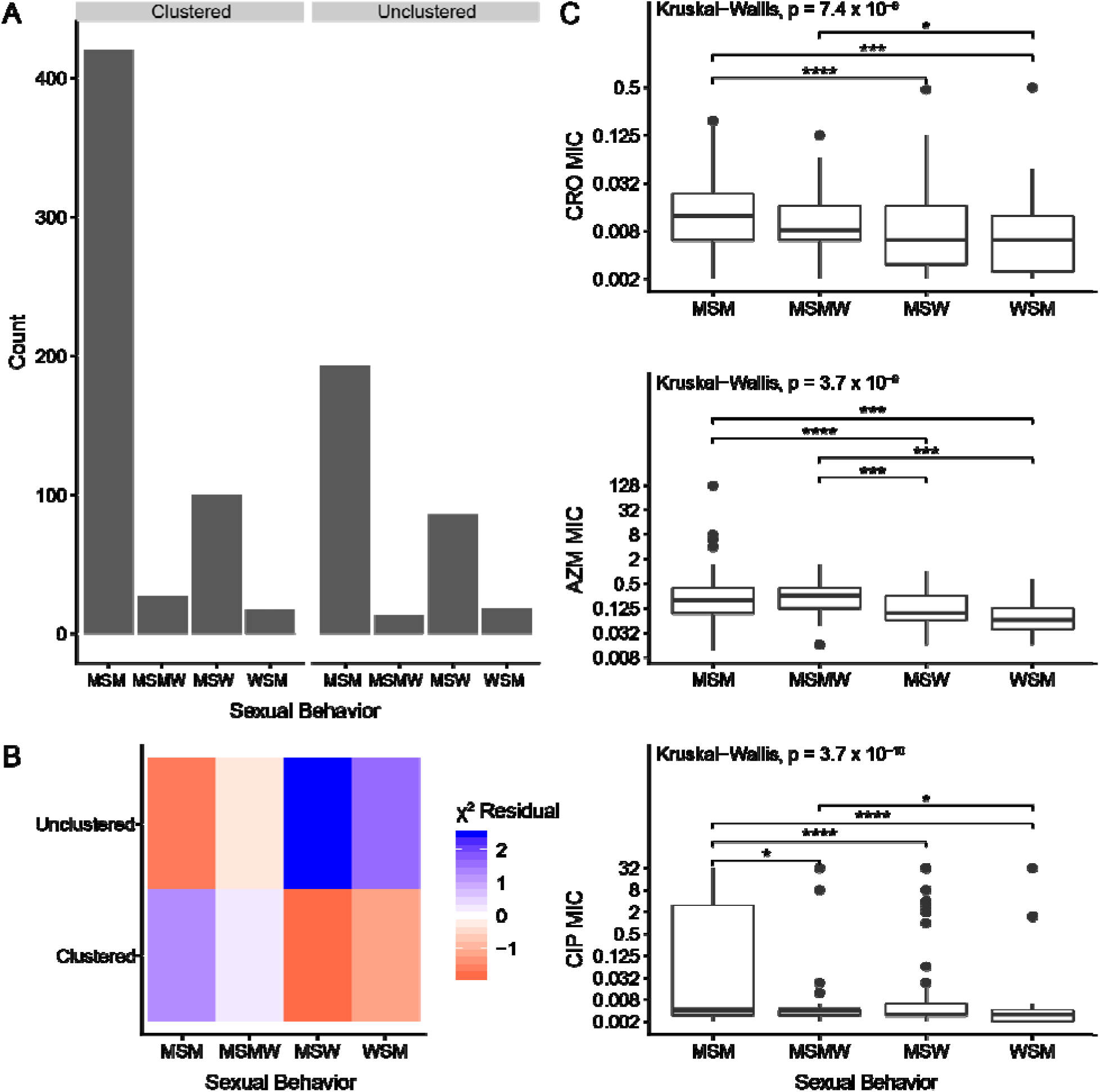
Sexual behavior groups are associated with clustering and antibiotic susceptibility. **A)** Sexual behavior groups and clustering are associated. Isolates were considered clustered if they were grouped with at least one other isolate in the dataset using the 10 non-recombinant SNP cutoff. We identified a significant association between sexual behavior groups and whether or not an isolate was clustered (p = 5.0 × 10^−4^). **B)** Clustered isolates are associated with MSM, and unclustered isolates are associated with heterosexuals. The χ^2^ residual for each category is displayed where blue represents more isolates than expected for the category and red represents fewer isolates than expected for the category. **C)** MSM-associated isolates have higher MICs across antibiotics. Significant pairwise comparisons are denoted by a bracket and asterisks (^*^ p < 0.05, ^**^ p < 0.01, ^***^ p < 0.001, ^****^ p < 0.0001).

**Figure 3.**
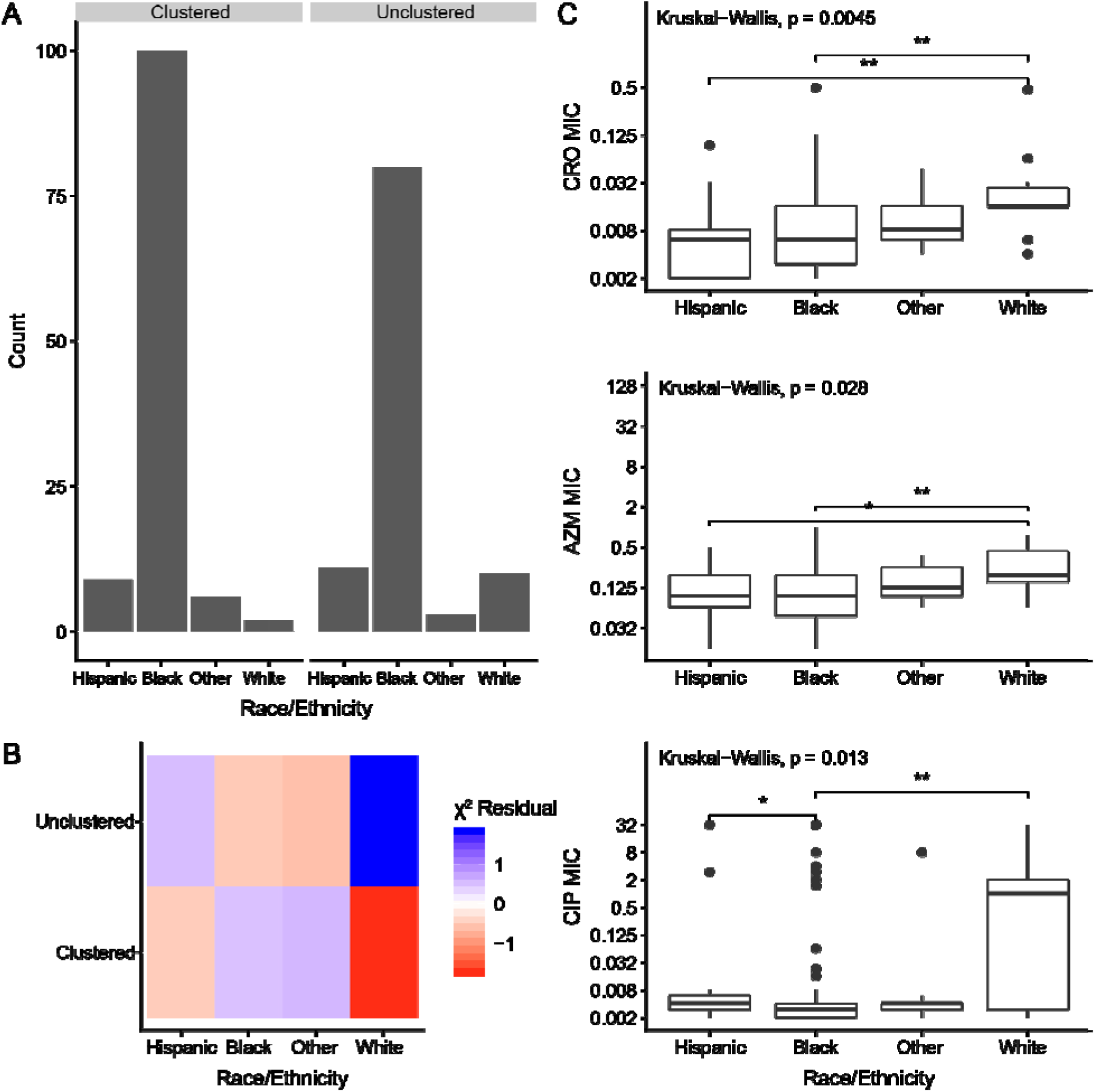
Race/ethnicity is associated with clustering and antibiotic susceptibility among heterosexuals. **A)** Race/ethnicity of heterosexuals and clustering are associated. Isolates were considered clustered if they were grouped with at least one other isolate in the dataset using the 10 non-recombinant SNP cutoff. We identified a significant association between heterosexual race/ethnicity and whether or not an isolate was clustered (p = 0.04563). **B)** Unclustered isolates are associated with white heterosexuals. The χ^2^ residual for each category is displayed where blue represents more isolates than expected for the category and red represents fewer isolates than expected for the category. **C)** Isolates from white heterosexuals have higher MICs than isolates from black heterosexuals across antibiotics. Significant pairwise comparisons are denoted by a bracket and asterisks (^*^ p < 0.05, ^**^ p < 0.01).

### Antibiotic susceptibility

Among isolates in our sample that underwent antibiotic susceptibility testing at NYC PHL, 24.3% (216/889) were resistant to ciprofloxacin, 0.9% (8/889) had reduced susceptibility to azithromycin, 0.3% (3/887) had reduced susceptibility to ceftriaxone, and 0.1% (1/886) had reduced susceptibility to cefixime. No isolates had reduced susceptibility to both ceftriaxone and azithromycin. The largest clusters were susceptible to currently recommended empiric therapy (ceftriaxone and azithromycin) as well as cefixime and ciprofloxacin (Figure 4). Within clusters, we observed isolates with a wide range of MICs. For some clusters (27, 79, 2, 5), this range is attributable a 2 bp deletion disrupting the reading frame of *mtrC*, a component of the Mtr efflux pump in a subset of isolates.

**Figure 4.**
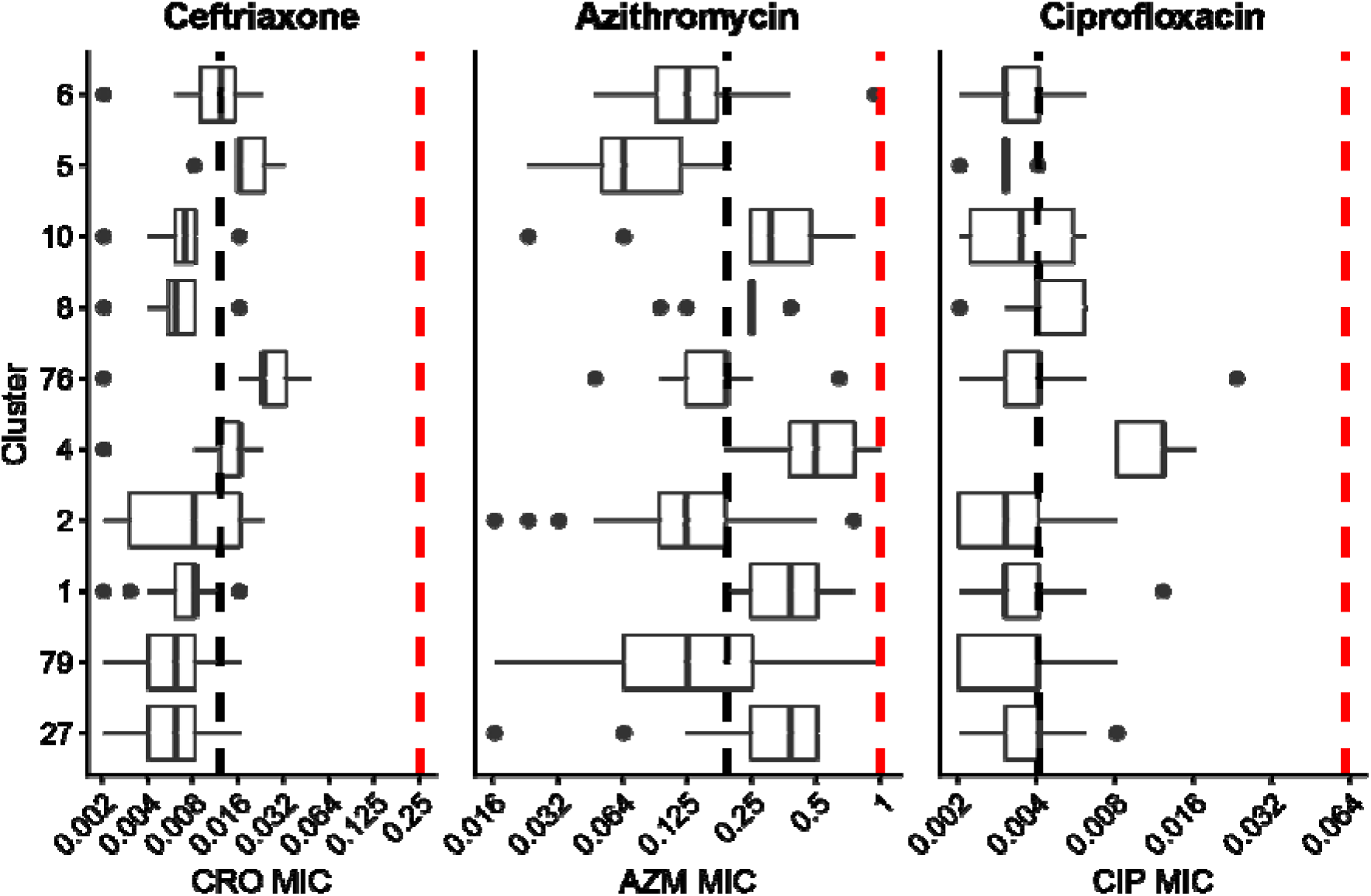
Isolates from large transmission clusters are sensitive to antibiotics. Minimum inhibitory concentrations (MIC, μg/mL) to ceftriaxone (CRO), azithromycin (AZM), and ciprofloxacin (CIP) for each isolate were measured using E-test. Cluster numbers correspond to Figure 1C. Black dashed lines represent median MICs for the dataset. All isolates have MICs below the susceptibility breakpoint (red dashed line) for all three antibiotics (CRO <= 0.25 μg/mL, AZM <= 1 μg/mL, CIP <= 0.06 μg/mL). Isolates in clusters do not have uniformly elevated MICs compared to the overall distribution, suggesting that antibiotic resistance was not the main driver of gonorrhea transmission in NYC during this time period.

As defining the patterns of the spread of isolates exhibiting antibiotic resistance may aid in surveillance and intervention strategies, we examined associations between MICs and patient demographics. Isolates from MSM had significantly higher mean MICs than isolates from heterosexuals (Figure 2C). However, no significant differences persist after controlling for lineage for ceftriaxone and azithromycin; ciprofloxacin MICs are significantly higher in lineage A isolates from MSM (p = 0.004, Supplementary Figure 3).

Among isolates from MSM, we found no differences across race/ethnicity subgroups in the mean MICs of ceftriaxone and azithromycin; however, there was an association between MSM race/ethnicity and ciprofloxacin MICs (p = 0.028; Supplementary Figure 4), with greater resistance in isolates from white MSM. In heterosexuals, we found significant associations between race/ethnicity and mean MICs for ceftriaxone, azithromycin, and ciprofloxacin (Figure 3C), with MICs for isolates from white heterosexual patients significantly higher than in isolates from black heterosexual patients (Figure 3C). These differences may also be attributable to lineage: of the 12 isolates from white heterosexual patients, only one was in lineage B.

## Discussion

We incorporated the analysis of phenotypic susceptibility results, patient demographics, and gonococcal genome sequences to investigate transmission in a major metropolitan area.. We found that the NYC population contains representatives from most major groups in a global sample. We also found that *N. gonorrhoeae* lineages were associated with sexual behavior groups, and we found a significant association between antibiotic susceptibility and patient demographic groups. We identified transmission clusters and found that the largest clusters were associated with patients from multiple demographic groups and consisted of *N. gonorrhoeae* isolates susceptible to gonorrhea therapy.

Our dataset comprised a retrospective sample of isolates derived from specimens collected from patients presenting at NYC SHCs for surveillance, diagnosis, and treatment of gonorrhea. Other providers (private providers, family planning, and others) make most of the reported gonorrhea diagnoses in NYC (82%), and our results show that not all patient populations are equally likely to be sampled for culture and susceptibility testing at SHCs. A large portion of NYC isolates (~35%) in our dataset were not clustered with any other isolate, suggesting that these transmission networks were not well sampled. Transmission links may have been missed in the setting of mixed infection because we sequenced single colony isolates. Isolates from MSM were more likely to be clustered than isolates from heterosexuals, and among heterosexuals, isolates from non-white patients were more likely to be clustered. This is likely related to sampling at SHCs, both because of the populations that attend SHCs [31] and the criteria for sending a specimen for culture, but may also be due to differences in sexual behavior.

Despite the convenience sampling, several findings emerge from our study. We found that almost all major clades from a global collection of *N. gonorrhoeae* genomes were represented in the NYC sample, highlighting the frequent transmission between regions and lack of geographic structure, consistent with other studies [9,32–34]. The phylogeny of the NYC *N. gonorrhoeae* population can be divided into the previously described two major global lineages, lineages A and B. A previous study of a global collection found a significant association between isolates from female patients and lineage B driving a hypothesis that isolates from lineage B are primarily found in heterosexuals and isolates from lineage A in MSM [8]. In our dataset, we confirmed these associations between lineage and sexual behavior groups in the setting of a metropolitan area. However, our data also suggested bridging between sexual behavior groups in the largest transmission clusters. One possible explanation for this seemingly contradictory result is that frequent bridging is a recent phenomenon, and the phylogenetic structure is the result of historical sexual networks. Another possibility is that while frequent bridging occurs, lineage A isolates are more successful in MSM networks and lineage B isolates are more successful in heterosexual networks. Further studies are needed to understand the relationship between *N. gonorrhoeae* population structure and adaptation to different sexual behavior groups.

Studies analyzing HIV status in the context of gonorrhea transmission networks in other regions have observed a lack of serosorting among HIV positive and HIV negative individuals [10,35,36]. As HIV pre-exposure prophylaxis (PrEP) was uncommon during 2012-2013 [37,38], our observations of gonorrhea transmission clusters with both HIV positive and negative patients suggest that in the sample population from NYC effective serosorting was not practiced in the era prior to PrEP introduction. Our results support CDC guidelines that a diagnosis with gonorrhea is an indication for PrEP [39].

Isolates from MSM had higher mean MICs across antibiotics compared to isolates from heterosexuals, primarily due to differences in susceptibility in lineages A and B. Data from national surveillance (GISP) also showed an association between MSM-associated isolates and reduced susceptibility to antibiotics: in 2018, 8.2% of isolates from MSM had azithromycin MICs ≥2.0 μg/mL compared to 2.4% of isolates from MSW, and 0.22% of isolates from MSM had ceftriaxone MICs ≥0.125 μg/mL compared to 0.16% of isolates from MSW [1]. Resistant *N. gonorrhoeae* strains may spread faster in MSM populations compared to heterosexual populations because of higher treatment rates among MSM [40].

We also found that among heterosexuals, isolates from white patients had higher MICs than isolates from other races/ethnicities, and the majority of isolates from white heterosexual patients belonged to lineage A. This may reflect differences in overall antibiotic use, diagnosis, or treatment in these populations. A surveillance study of quinolone resistant *N. gonorrhoeae* in southern California in 2001-2002 found that non-white patients were at lower risk for quinolone resistant gonorrhea [41]. There is a relationship between seasonal azithromycin use and resistance in *N. gonorrhoeae*, suggesting a role for bystander selection (selection for resistance in bacteria other than the intended target of therapy) in the development and maintenance of resistance in the *N. gonorrhoeae* population [42]. Given that white Americans may consume twice the antibiotic prescriptions compared to other race/ethnicities [43,44], *N. gonorrhoeae* infecting white Americans may be subject to increased bystander selection. More studies on the relationship between antibiotic use across demographic groups and their risk for resistant gonorrhea are needed.

In the United States, genomics has been primarily used to understand the biology and transmission of antibiotic resistant gonococcus. However, the major transmission clusters in our study involved strains that were susceptible to currently recommended empiric therapy, as well as cefixime and ciprofloxacin, indicating that during the study period antibiotic resistance was not a major driver of gonorrhea transmission in NYC. This dataset was collected primarily in 2012-2013, and resistance to azithromycin may have contributed to more recent gonorrhea outbreaks [13], as azithromycin reduced susceptibility has increased from 0.6% to 4.6% from 2013 to 2018 [1]. Within clusters, we observed a wide range of MICs. In some clusters, all with evidence of bridging, the range of MICs could be attributed to a portion of isolates encoding a 2bp deletion in *mtrC*, which is associated with antibiotic susceptibility and the cervical niche [30]. While resistance remains a major public health concern, strategies to reduce overall gonorrhea transmission are also needed as pre-existing transmission networks may present opportunities for rapid spread of resistant lineages. Additionally, fewer cases overall allows public health programs to concentrate more resources on resistant cases. Greater understanding of the transmission dynamics of both susceptible and resistant infections can aid the design of effective intervention strategies for controlling gonorrhea, and further investment in sexual health services and interventions are critical.

## Data Availability

Whole genome sequencing data were deposited in the European Nucleotide Archive (ERA) under study accession PRJEB10016. All additional data and scripts are available at https://github.com/gradlab/GC_NYC.

https://www.ebi.ac.uk/ena/data/view/PRJEB10016

https://github.com/gradlab/GC_NYC

## Funding

This work was supported by the National Institute of Allergy and Infectious Diseases at the National Institutes of Health [R01 AI132606 and 1 F32 AI145157-01] and the Wellcome Trust [098051].

## Acknowledgements

Portions of this research were conducted on the O2 High Performance Compute Cluster, supported by the Research Computing Group, at Harvard Medical School. The authors thank Greicy Zayas, Inessa Rubenstein, Evelyn Patricio, Angela Liang, Sabine Glaesker, Bun Tha, and Teresa Rozza from the NYC DOHMH PHL. The authors additionally thank Kevin Ma and Allison Hicks for their contributions to the global dataset. We also thank Deborah Williamson for her comments on the manuscript.

## Supplementary Text

### Definining transmission clusters

We did not have information on sex partner links in our patient population, so we could not define a SNP distance cutoff consistent with sexual partnerships. While a 12 SNP cutoff, proposed based on time scaled phylogenies [33], is consistent with the median time between collection dates of our samples (7.7 months), other studies of transmission have used a cutoff of 10 SNPs to identify probable transmission events based on SNP differences between named partners [10]. On clustering isolates based on the more conservative SNP distance (10 SNPs), we observed that isolates nested within a cluster on the phylogeny were not included in the cluster using our method because the genetic distances between the isolate and other members of the cluster were greater than the SNP cutoff (Supplementary Figure 1). One explanation for increased SNP distances could be that the specimen from which the isolate was cultured was collected at a different time compared to other isolates in the cluster, and therefore, presumably a greater number of mutations had accumulated than our SNP cutoff. However, we found that 74% of nested isolates or clusters had specimen collection dates within the range of the cluster. Manual inspection of these isolates showed that the genomes contained unique recombination events identified by Gubbins [24]; the median number of unique recombination blocks for singleton isolates nested within a cluster was 5 (IQR: 0.25-9), compared to a median of 0 (IQR: 0-3) for the entire dataset. We observed SNPs at the boundaries of these events or additional clusters of homoplastic SNPs not identified as recombinant, which was inflating SNP distances from the rest of the cluster (Supplementary Figure 1). Thus, we included any isolate descended from the most recent common ancestor (MRCA) of a cluster for final clustering (Supplementary Figure 2). Clustering with a 12 SNP cutoff did not substantially change the results of clustering analyses (Supplementary Table 3).

**Supplementary Figure 1.**
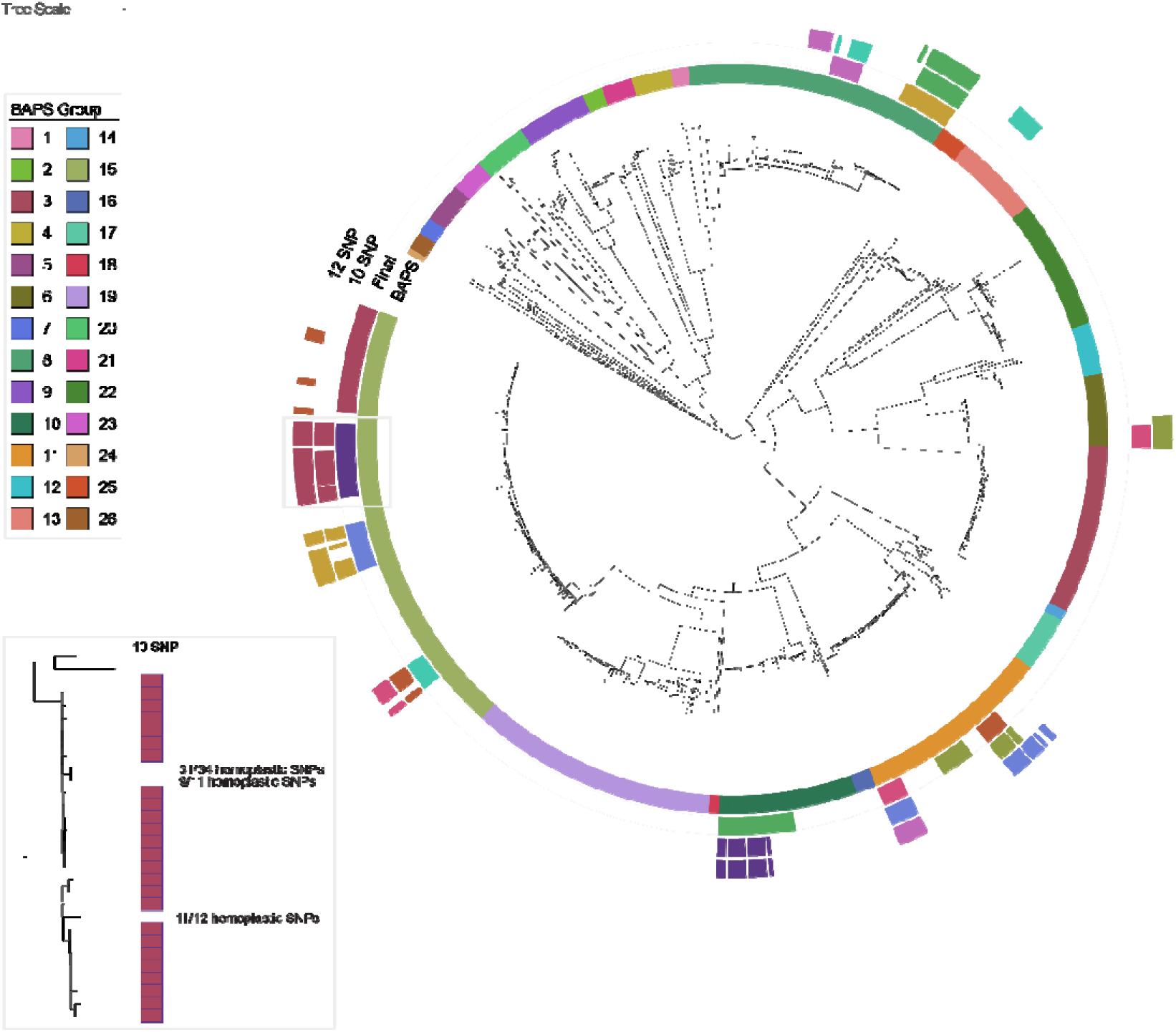
Clustering of *N. gonorrhoeae* isolates from New York City. The innermost annotation ring corresponds to clusters identified using a phylogeny-based analysis in fastbaps. The second annotation ring corresponds to the 10 largest final transmission clusters defined using a 10 non-recombinant SNP cutoff with any additional nested isolates. The third and fourth annotation rings correspond to the 10 largest transmission clusters using 10 non-recombinant SNP and 12 non-recombinant SNP cutoffs, respectively. Without correcting for undetected recombination (see inset), transmission clusters were missing nested isolates. **Inset**. A portion of the phylogenetic tree corresponding to final cluster 1 (dark purple in Supplementary Figure 1 annotation ring) is shown on the left. Black boxes correspond to isolates considered part of the cluster using a 10 non-recombinant SNP cutoff. Isolates nested within the phylogeny but with distances greater than 10 SNPs have homoplastic SNPs contributing to their distance, which may represent undetected recombination events.

**Supplementary Figure 2.**
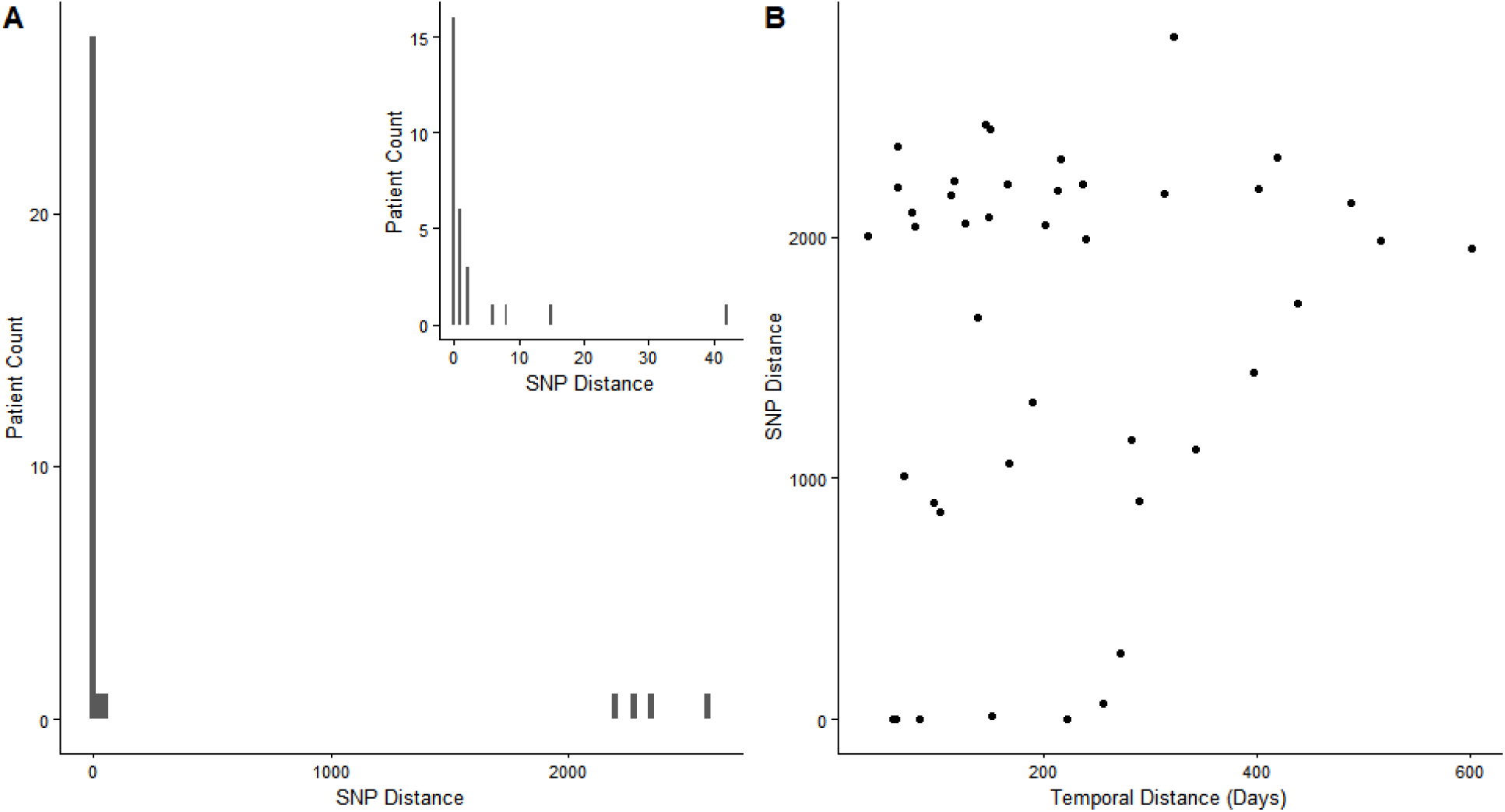
Genetic distance between isolates in patients with multiple samples. **A)** Patients with multiple samples collected at the same visit can be infected with multiple strains. The non-recombinant SNP distance between isolates collected on the same day is shown on the X axis, and the number of patients is shown on the Y axis. 18.2% of patients are infected by multiple strains. **B)** Patients who return to the clinic are infected by new strains. The temporal distance between clinic visits is plotted on the X axis, and the non-recombinant SNP distance between isolates is plotted on the Y axis. 90.5% of observed reinfections were with a new strain.

**Supplementary Figure 3.**
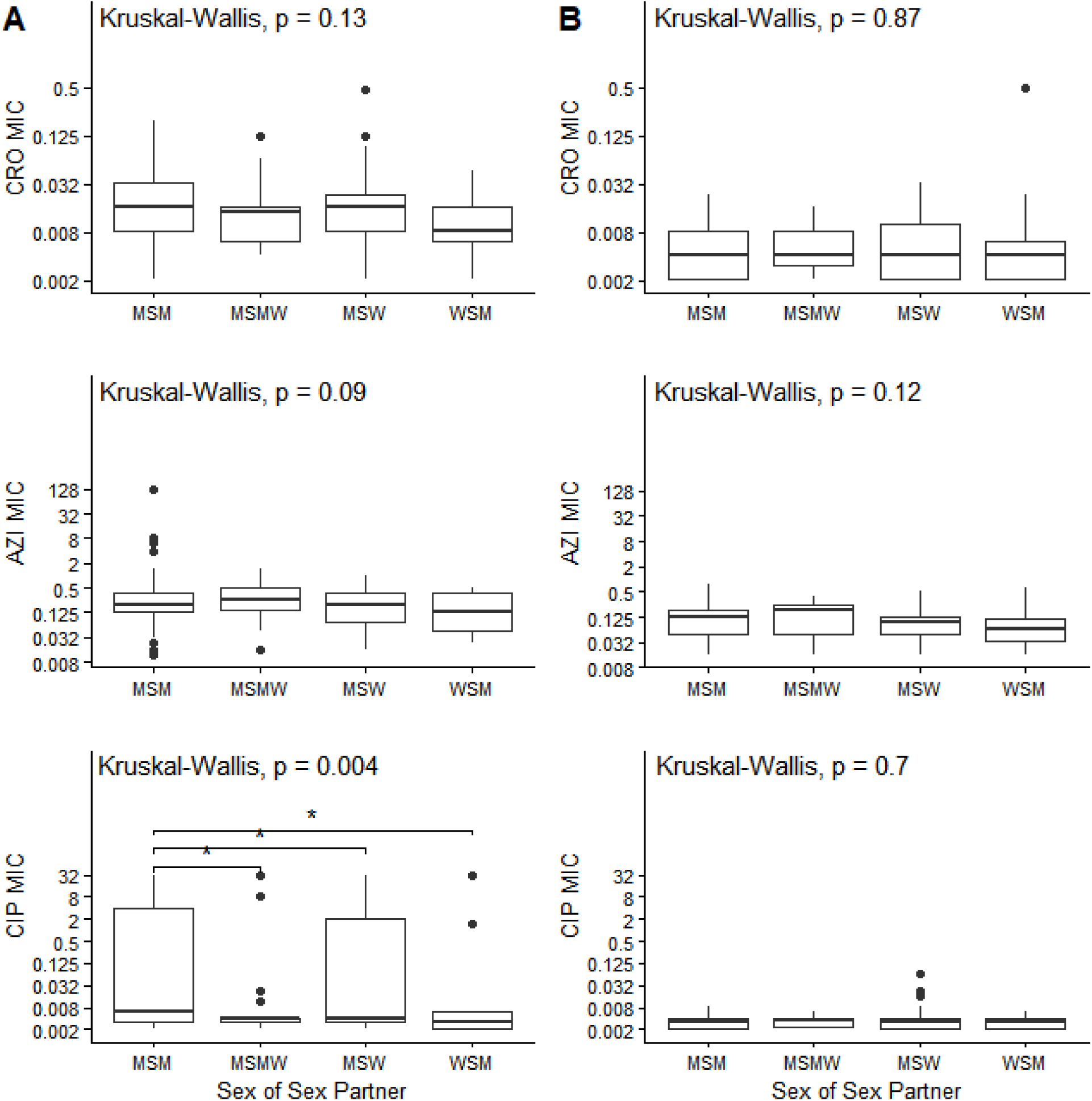
The association between MICs and sexual behavior is explained by lineage differences for ceftriaxone and azithromycin. A) Lineage A B) Lineage B Significant pairwise comparisons are denoted by a bracket and asterisks (^*^ p < 0.05).

**Supplementary Figure 4.**
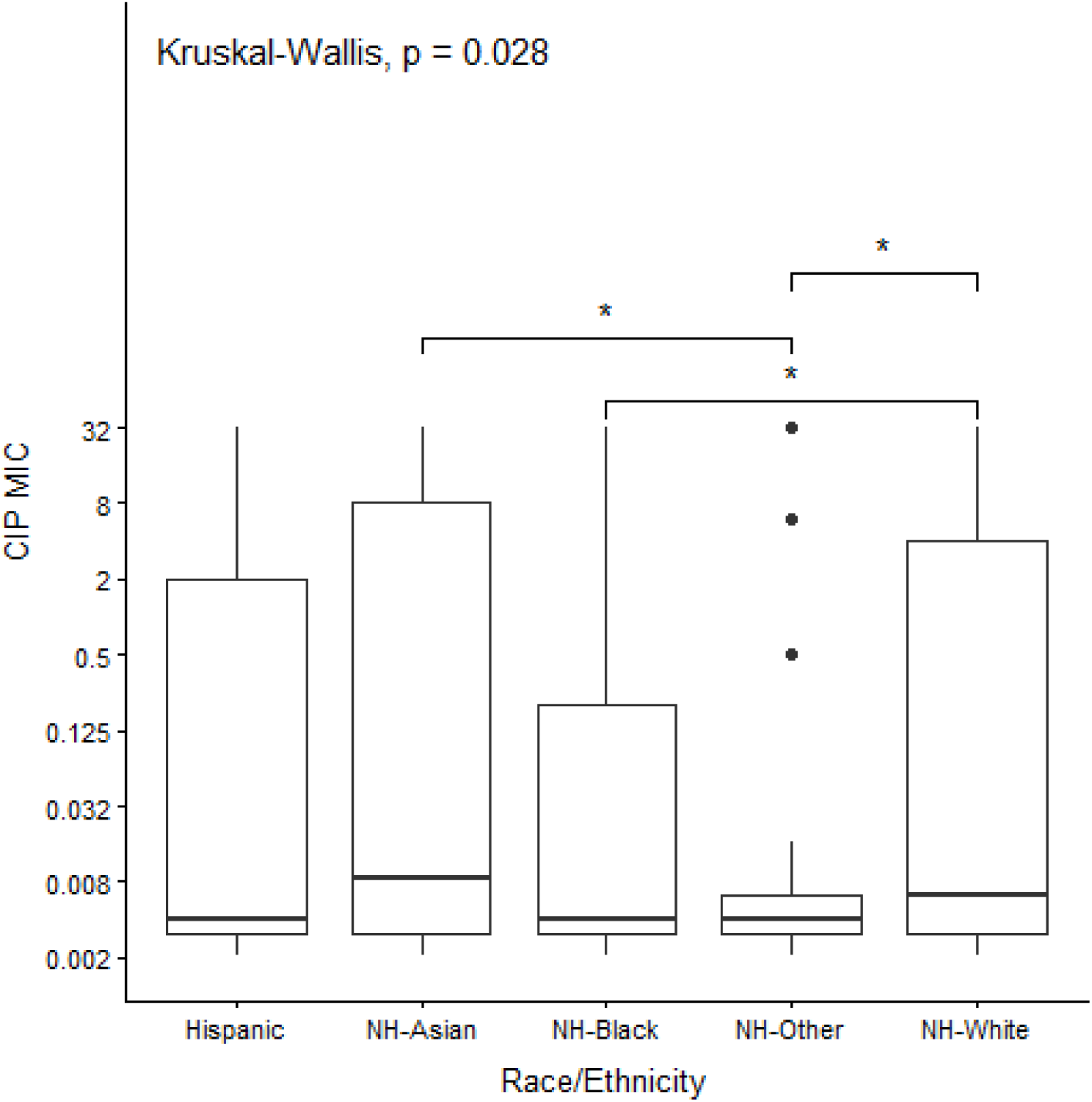
MSM race/ethnicity is associated with ciprofloxacin MICs. Significant pairwise comparisons are denoted by a bracket and asterisks (^*^ p < 0.05).

**Supplementary Table 1**. NYC isolate accessions and metadata.

**Supplementary Table 2**. Global dataset accessions and BAPS groups.

**Supplementary Table 3**. Clustering of isolates with 12 SNP cutoff and addition of nested isolates.

| Observation from 10 SNP cutoff | Value with 12 SNP cutoff |
| --- | --- |
| Clusters $\geq 10$ samples show evidence of bridging | 8/13 large clusters with evidence of bridging |
| Association between sexual behavior groups and clustering | p-value = 0.0003278 |
| Association between heterosexual race/ethnicity and clustering | p-value = 0.01723 |

## Notes

### Competing Interest Statement

The authors have declared no competing interest.

